# Acquired decrease of the C3b/C4b receptor (CR1, CD35) and increased C4d deposits on Erythrocytes from ICU COVID-19 Patients

**DOI:** 10.1101/2020.08.10.20162412

**Authors:** Aymric Kisserli, Nathalie Schneider, Sandra Audonnet, Thierry Tabary, Antoine Goury, Joel Cousson, Rachid Mahmoudi, Firouze Bani-Sadr, Lukshe Kanagaratnam, Damien Jolly, Jacques HM Cohen

## Abstract

In order to study the mechanisms of COVID-19 damage following the complement activation phase occurring during the innate immune response to SARS-CoV-2, the CR1, receptor regulating complement activation factor, CR1 (CD35, the C3b/C4b receptor), C4d deposits on Erythrocytes (E), and the products of complement activation C3b/C3bi were assessed in 52 COVID-19 patients undergoing O_2_ therapy or assisted ventilation in ICU units in Rheims France.

An acquired decrease of CR1 density of E from COVID-19 patients was observed (Mean 418, SD 162, N=52) versus healthy individuals (Mean = 592, SD = 287, N= 400), Student’s t-test p<10^−6^, particularly among fatal cases, and paralleling several clinical severity parameters.

Large deposits of C4d on E in patients were well above values observed in normal individuals, mostly without concomitant C3 deposits, in more than 80% of the patients. This finding is reminiscent of the increased C4d deposits on E previously observed to correlate with sub endothelial pericapillary deposits in organ transplant rejection, and with clinical SLE flares. Conversely, significant C3 deposits on E were only observed among ¼ of the patients.

The decrease of CR1/E density, deposits of C4 fragments on E and previously reported detection of virus spikes or C3 on E among COVID-19 patients, suggest that the handling and clearance of immune complex or complement fragment coated cell debris may play an important role in the pathophysiology of SARS-CoV-2. Measurement of C4d deposits on E might represent a surrogate marker for assessing inflammation and complement activation occurring in organ capillaries and CR1/E decrease might represent a cumulative index of complement activation in COVID-19 patients.

Taken together, these original findings highlight the participation of complement regulatory proteins and indicate that E are important in immune pathophysiology of COVID-19 patients. Besides a potential role for monitoring the course of disease, these observations suggest that novel therapies such as the use of CR1, or CR1-like molecules with the aim of down regulating complement activation and inflammation for therapy should be considered.

**HIGHLIGHTS:**

- Acquired decrease of CR1 density on E in COVID-19 patients correlated with clinical severity and mortality.
- Large C4d deposits were found on E in most patients, reminiscent of those observed on E of patients undergoing organ transplant rejection that were associated with peri-capillary deposits, as well as on E from patients undergoing SLE flares.
- C4d deposits on E may be useful as a surrogate marker for inflammation and complement activation in organ capillaries.
- Decreased CR1/E may be useful as a cumulative index of complement activation in COVID-19 patients.
- The use of CR1 or CR1-like molecules for down-regulating complement activation for therapy should also be considered.
- These original findings indicate the participation of complement regulatory proteins in COVID-19 and on the role of E in immune mechanisms of the disease.

## 1. Introduction

We looked at the role of complement on erythrocytes (E) in hospitalized patients with COVID-19 to clarify the role for specific complement factors in the pathophysiology of the disease that may be used to monitor patients and suggest novel therapies.

Discovered in 1895 (Cavaillon et al., 2019), complement is a system of plasma and cyto-membrane proteins with three main functions: the labeling of certain microorganisms and antigens, promoting their attachment to the surface of cells carrying complement membrane receptors, and the attraction of phagocytes where there is complement activation and to promote cell lysis. There are three pathways to complement activation: the classic pathway, the alternate pathway and the lectin pathway (Edberg et al., 1987). The classic pathway requires prior specific recognition of an antigen by an antibody, while the other two are activated directly by the membranes of the pathogens. Complement, a system widely conserved during evolution, owes its name to the observation that it acts in addition to (“complementing”) antibodies in the lysis of bacteria. It was later discovered that complement is also important in innate immune defense, as well as the main scavenger of dead cells or damaged tissue. As a component of the innate immune response, it can be mobilized immediately, and not specifically, for a given antigen. On the other hand, complement is involved in the initiation and progression of the adaptive immune response (Walport et al., 2001).

The activation of the various components of complement takes place as a cascade, in a manner similar to that observed during coagulation or fibrinolysis. Some of the components endowed with enzymatic activity circulate in an inactive form, acquiring their proteolytic or biological activity only after limited proteolysis, thereby becoming the activator of the next protein in the activation cascade, so each step is amplified. Besides this cascade amplification, the consequence of complement protein activation is the appearance of various biologically active cleavage products capable of interacting with many cell types via specific receptors. Thus, for example C5a, C3a and C4a have anaphylatoxic activity via receptors located on monocytes-macrophages, mast cells and polymorphonuclear cells (Murakami et al., 1993; Karapesi et al., 2006; Sacks et al., 2003).

C4d is the product of the cleavage of C4b, which is the product of the cleavage of C4 (Atkinson, 1988) during activation of C4 by the C1q, r, s complex (the classical pathway) and by the mannan-binding lectin (MBL) – mannan binding lectin serine protease (MASP) 1 and 2 complex (the lectin pathway). Additional mechanisms of direct activation of C4, like C3 activation, have been suspected but remain uncertain. The 42 KDa single chain C4d is a short covalently bound cleavage product of the C4 molecule. Most of the polymorphisms of C4are located in the C4d fragment (Atkinson et al., 1988).

C4d has been known since the 1970s as the biochemical basis of the Chido/Rodgers Blood Group (Tilley et al., 1978). It covalently binds to cell surfaces or immune-complexes via a thio-ester bridge for an extended period of days to months (Van Del Elsen et al., 2002). C4d can be detected on the surface of E in all mammals under physiologic conditions (Ferreira, 1980). The origin of C4d on E surface is not known. One hypothesis is that, like C3d, the C4 molecule self-activates at a low, steady state, like a night light ready to light up fully when needed. This phenomenon of a system running at idle is described by the term “tick over” (Atkinson et al., 1988).

The detection of E C4d has been performed by flow cytometry in healthy subjects in a reproducible manner since the early 1990s (Freysdottir and Sigfusson, 1991). Covalent C4d deposits on E are present at low levels in normal individuals. The level of E C4d is increased in pathologic states involving circulating immune complexes, such as autoimmune hemolytic anemia, autoimmune throm-bocytopenia (Leddy et al., 1974) and immuno-allergic reactions (Giles and Robson, 1991). Higher densities of C4d on E have been found in SLE at the time of flares of the disease (Manzi et al., 2004), as well as in kidney transplant rejection (Golocheikine et al., 2010; Haidar et al., 2012), where it was associated with the sub-endothelial capillary C4d deposits commonly found in chronic vascular rejection, without C3 or Ig deposits in most cases (Feucht et al., 1991). C4d deposition in organs has also been observed in the rejection of other organ transplants (Lee et al., 2008). Most importantly, C4d deposits in lung capillaries have been reported (Magro et al., 2020) in COVID-19 patients.

The complement receptor type 1 (CR1, CD35) regulates the activity of complement. By interfering with its ligands C3b, C4b, C3bi (Fearon, 1980; Dobson at al., 1981; Schreiber et al., 1981; Ross et al., 1983), a subunit of the first complement component, C1q (Klickstein et al.; 1987) and MBL (Ghiran et al., 2000), CR1 inhibits the activation of complement (Iida and Nussenzweig, 1981) and is involved in the humoral and cellular immune responses. CR1 is a transmembrane glycoprotein present on the surface of many cell types, such as E (Fearon, 1980), B lymphocytes (Ross et al., 1978), some T cells, cells of the monocytic line, follicular dendritic cells (Reynes et al., 1985), glomerular podocytes (Pascual et al., 1994) and fetal astrocytes (Gasque et al., 1996). CR1 on E in humans binds with and carries immune complexes or cell fragments to spleen or liver for their clearance from the blood, (Cornacoff et al., 1983; Waxman et al., 1984; Waxman et al., 1986), while most other mammals use platelets. CR1 also contributes to the inactivation of the C3 amplification loop also known as alternative pathway, as a co-factor of Factor I (Inactivator).

CR1/E density varies from one individual to another (from 100 to 1200 CR1 / E), and from one E to another in the same individual. Despite that low density, the number of E makes CR1/E the major source of CR1 available in the blood. Some very rare individuals with the “null” KN phenotype express fewer than 20 CR1/E (Pham et al., 2010). The density of CR1/E is regulated by two co-dominant autosomal alleles linked to a point mutation in intron 27 of the gene coding for CR1 * 1 (Wilson et al., 1986; Rodriguez De Cordoba and Rubinstein, 1986). This mutation produces an additional restriction site for the *Hin*dIII enzyme. The restriction fragments obtained after digestion with *Hin*dIII in this case are 7.4 kb for the allele linked to a strong expression of CR1 (H: High allele) and 6.9 kb for the allele linked to low CR1 expression (L: Low allele). This link is always found in most populations except a single African tribe (Herrera et al., 1998). The level of expression of E CR1 is also correlated with the presence of point nucleotide mutations in exon 13 encoding SCR 10 (I643T), in exon 19 encoding SCR16 (Q981H). It is high in homozygous (643I / 981Q) and low in homozygous individuals (643T / 981H) (Birmingham et al., 2003). Thus “low” individuals express around 150-200 CR1 / E, “medium” individuals express around 500-600 CR1 / E and “high” individuals express around 800-1000 CR1 / E. In addition to this E density polymorphism, CR1 is also characterized by antigenic polymorphisms corresponding to the blood group KN (Moulds et al., 1992) and a length polymorphism corresponding to 4 allotypes of different sizes: CR1*1 (190 KDa), CR1*2 (220 KDa), CR1*3 (160 KDa) and CR1*4 (250 KDa) (Dykman et al.,1985).

CR1/E decreases by one third during the 120-day life of E in normal individuals (Cohen et al., 1992), but a larger proportion of CR1/E can be lost in acute situations, making CR1/E measurement a cumulative assessment of what happened during the E lifespan, like HbA1c, but in the opposite direction. An acquired decrease of CR1/E has been described in AIDS (Jouvin et al., 1987), SLE (Cohen et al., 1992), Malaria (Waitumbi et al., 2004) and Alzheimer disease (Mahmoudi et al, 2015; Mahmoudi et al, 2018).

One paper reported on a decrease of CR1/E in patients during the SARS epidemic (Wang et al., 2005). We hypothesized that the major clinical crisis observed in COVID-19 disease might be induced by the immune response and could lead to a major consumption of CR1 on E. Evidence is accumulating on the role of the complement system in SARS (Gralinski et al., 2018) and now in COVID-19 (Risitano et al., 2020; Magro et al., 2020; Holter et al., 2020, Carvelli et al., 2020), (Berzuini et al., 2020) as well as on immune-adherence on E in that disease (Metthew Lam et al., 2020).

Given these observations we chose to look at the CR1/E antigenic site, C3b/C3bi on E and C4d/E deposit measurements in patients with COVID-19.

## 2. Material and Methods

### 2.1 Patients and controls individuals

We performed an observational study in which we determined CR1/E density and the levels of C3b/C3bi and C4d deposits on E in 52 COVID-19 patients undergoing O_2_ therapy or assisted ventilation in ICU units in Rheims, France. The number of longitudinal samples from a given individual ranged from one to five, leading to 114 samples from 52 patients, 32 of the patients having been tested more than once (supplemental Fig. 1). The first determination of CR1/E, or the lowest CR1/E level reached during longitudinal follow-up CR1/E determination for a given patient, were used to perform comparisons to different clinical or biological parameters from which the lower or the higher recorded value were considered. (Lowest SaO2% or Highest D-Dimer value).

**Fig. 1.**
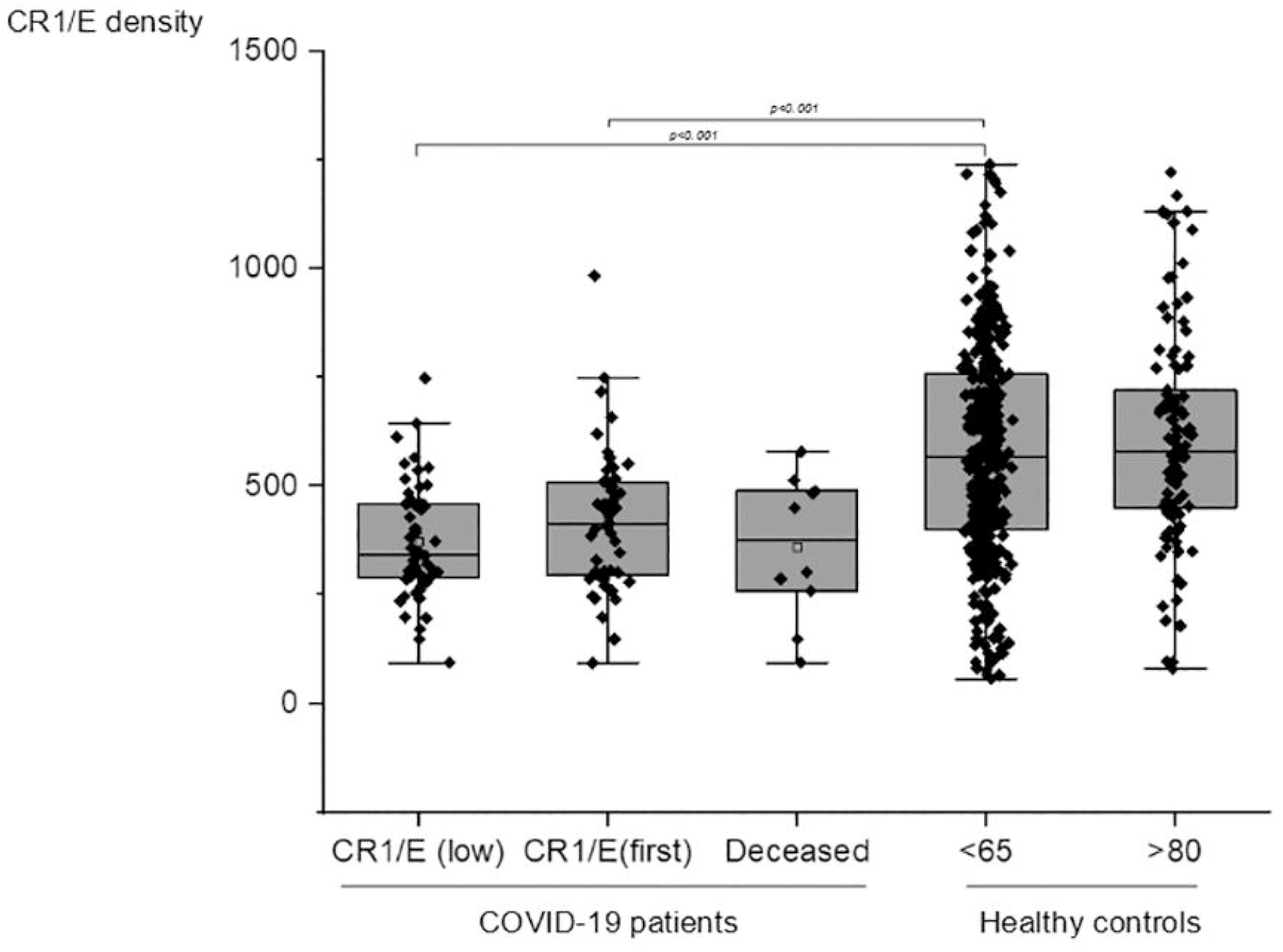
Comparison of CR1/E density between COVID-19 patients and healthy individuals. CR1 antigenic sites on E. CR1/E (low): from patients at the lowest value during longitudinal follow-up. (Mean = 372 SD = 134 N = 52). CR1/E (first): CR1 on erythrocyte from patients at the first determination (Mean = 418 SD = 162 N = 52). Deceased: CR1/E among patients deceased as in-patients (Mean = 361 SD = 165 N = 10). <65: CR1/E among healthy blood donors below 65 year old (Mean = 592 SD = 287 N = 400). >80: healthy controls above 80 years (Mean = 606 SD = 248 N = 98). Means are depicted as open squares. Patients at first determination, at the lowest determination during follow-up, or deceased, versus Healthy blood donors displayed a significant lower density of CR1/E. Wilcoxon test p<0.001).

Data from historical cohorts of healthy volunteers, 400 blood donors under 60 years and 98 healthy individuals above 80, were used as CR1/E control populations. In addition, E from 3 healthy individuals with known values of CR1/E and C4d/E were used to set-up calibration curves for CR1/E and as positive threshold for C3 or C4d/E deposits. An historical C4d healthy cohort of 88 individuals was used as a reference population (Haidar et al., 2012).

### 2.2 CR1/E antigenic site, C3b/E and C4d/E deposit measurements

We determined the CR1 density and the presence of C3b/C3bi and C4d deposits on E using flow cytometry. After being washed three times in Bovine Serum Albumin– Phosphate Buffered Saline (PBS/BSA) 0.15% buffer, E were incubated with biotinylated J3D3 anti-CR1 (C3b/C4b receptor CD35) monoclonal antibody (non-commercial monoclonal antibody, courtesy of Dr J. Cook) for 45 min at 4°C. Bound antibodies were revealed using an enhancing Phycoerythrin-Streptavidin (reference: 554061, BD Biosciences, San Jose, CA, USA)/ Vector anti-streptavidin biotin (reference: BA-0500, Eurobio Ingen, Les Ulis, France). Reference E of known CR1 densities allowed us to establish a calibration curve and absolute antigenic site number for a given sample (Cohen et al., 1987; Kisserli et al., 2020).

The deposits of C4d on E were evaluated following the same protocol but using a biotin conjugated monoclonal antibody (Biotinylated Anti-Human C4d (reference: A 704, Quidel, San Diego, CA, USA). The deposits of C3b and C3bi was evaluated using only a FITC conjugated monoclonal antibody (FITC Anti-Human complement component C3b/C3bi mAb; reference: CL7631F, Cedarlane, Hornby, Ontario, Canada).

C3b/C3bi or C4d deposits expressed as the number of events in an arbitrary window at the right of the control staining distribution were considered as positive and compared to values observed in normal individuals. Reference C3b or C4d coated E for calibrating flow cytometry and standardizing inter-assay repeatability as previously described (Haidar et al., 2012; Kerr and Stroud, 1979) were not available due to the particular circumstances of the study, with no possibility of ordering reagents and limited physical access to the lab. Two individuals from the historical cohort were tested again together with the present COVID-19 patients, allowing us to convert the scale of the historic cohort initially set to a reference standard unavailable for the present study due to the limitations imposed by the lock down in place to control the epidemic.

Readings of CR1, C3b/C3bi or C4d staining were done on a LSRFORTESSA flow cytometer (reference: 647788, BD Biosciences, San Jose, CA, USA).

### 2.3. DNA Extraction

DNA from 3-mL whole blood samples was isolated using the QIAamp DNA Blood Mini Kit (Qiagen, Courtaboeuf, France). The manufacturer’s instructions were followed according to the recommended protocol. Briefly, tubes containing 200 µL of EDTA whole blood, 200 µL of lysis buffer (containing guanidine hydrochloride), and 20 μL of proteinase K were mixed and incubated for 10 min at 56 °C. After this incubation, 200 µL of ethanol (>96%) was added and mixed, and the samples were applied to the QIAamp Spin Column. After 2 centrifugations with washing buffers, the DNA was finally eluted with 60 μL of elution buffer after incubation for one minute and centrifugation.

### 2.4. CR1/E Density Genetic Polymorphism Using HindIII PCR-RFLP

The CR1 density polymorphism on E was determined using PCR amplification and *Hin*dIII restriction enzyme digestion, as described previously (Cornillet et al., 1991). The PCR primers used were 5′–CCTTCAATGGAATGGTGCAT–3′ and 5′–CCCTTGTAAGGCAAGTCTGG–3′. PCR was performed on a MyCycler apparatus (Bio-Rad, Marnes-la-Coquette, France) using the following conditions: a final volume of 100 µL containing 2 µL of DNA solution (approximately 100–250 ng/µL), a 200-µM concentration of each deoxynucleoside triphosphate, a 0.5 mM concentration of each primer, 2.5 mM MgCl_2_, and 2.5 U of AmpliTaq Gold DNA Polymerases (ThermoFisher Scientific, Illkirch, France) in the buffer supplied by the manufacturer. The amplification conditions were as follows: 10 min at 94 °C, followed by 40 cycles of 1 min at 94 °C, 1 min at 61 °C, and 2 min at 72 °C, before being held for 10 min at 72 °C.

For RFLP determination, 30 µL of PCR product and 2 µL of *Hin*dIII were incubated in a final volume of 50 µL in the buffer supplied by the manufacturer at 37 °C for 2 h, followed by analysis on a 2% ethidium bromide gel.

Using this protocol, *Hin*dIII digestion did not cleave the PCR product (1.8 kb) from individuals who were homozygous for the CR1 high-density allele (HH). The 1.8 kb band was fully cleaved into two smaller bands of 1.3 and 0.5 kb in samples from individuals who were homozygous for the CR1 low-density allele (LL).

The *Hin*dIII polymorphism rs11118133, the main codominant bi-allelic SNP governing CR1 density on E was determined for every patient using PCRFLP (Wilson et al., 1986) with appropriate controls without any ambiguous allele assessment, allowing the conversion of the absolute CR1/E antigenic site phenotypes observed to percentages of expected values based on their genotype, termed here CR1/E%.

### 2.5 Data analysis

Statistical analyses were performed using BiostaTGV software http://biostatgv.sentiweb.fr/ (Institut Pierre Louis d’Epidémiologie et de Santé Publique (IPLESP), Paris, France), Calcstat software http://www.info.univ-angers.fr/~gh/wstat/calcstat.htm, G. Hunaut, Angers University Angers France; and Origin software (OriginLab, Northampton, Massachusetts, USA). When Student’s t-test was used, the normal distribution of the concerned population was checked. Non parametric Wilcoxon tests were used for too small or non-normal sub-populations.

## 3. Results

### 3.1 CR1/E density from COVID-19 patients is decreased

CR1/E as antigenic sites were measured in 52 COVID-19 patients undergoing O_2_ therapy or assisted ventilation in ICU units in Rheims, France. A large decrease in CR1/E antigenic sites among patients was observed when compared to an historical cohort of healthy blood donors. When COVID-19 patients (Mean = 418 SD = 162 N = 52) were compared to healthy subjects (Mean = 592 SD = 287 N = 400), the mean density of E CR1 was significantly lower in patients (Student t-test p<0.001 (Fig. 1). As when considering the lowest CR1/E value observed for a given patient (Mean = 372 SD = 134 N = 52). Among the 10 deceased patients, a trend toward even lower CR1/E values was detected (Mean = 361 SD = 165 N = 10) (Fig. 1),

We also ruled out an age effect (mean age of the COVID-19 patients was 64) by comparing results of E CR1 density from blood donors to historical controls consisting of individuals over 80 years of age. Values for E CR1 density from the elderly controls (Mean = 606 SD = 248 N = 98) were similar to those for blood donors under 65 (Mean = 592 SD = 287 N = 400) (Fig. 1).

Some longitudinal data was available for 32 patients. We analyzed the dynamics of CR1/E over the course of the disease using the onset of symptoms as a proxy for the onset of the disease. Although most patients exhibited low CR1/E values early in the disease, a clear progressive decrease of CR1/E occurred around day 10 of symptoms in some individuals, whereas on follow-up 2-3 weeks later during recovery some individuals displayed a return to normal values in the normal range] (supplemental Fig. 1). Interestingly, 6 patients received blood transfusions before testing that didn’t dramatically increase CR1/E in active disease, even after one individual received 8 E packs. These observations suggest that the CR1/E of transfused E had been rapidly consumed as part of the disease process.

We looked at whether the acquired decrease in CR1/E density correlated with clinical severity parameters (supplemental Fig. 2 and supplemental Fig. 3). Forty-six patients with arterial O2 saturation less than 91% had a mean of E CR1 density that was lower than those with arterial saturation greater than 91% (49 % SD = 10 N = 14 versus 62 % SD = 10 N = 32; t-test with Welch correction p= 0.015) (Fig. 7). There was a trend toward a decreased density of CR1/E with higher levels of D-dimers. Higher levels of D-dimers indicate an increased level of coagulation. This data suggests that decreased CR1/E may reflect increased coagulation and thrombosis in COVID-19 patients (supplemental Fig. 3).

**Fig. 2.**
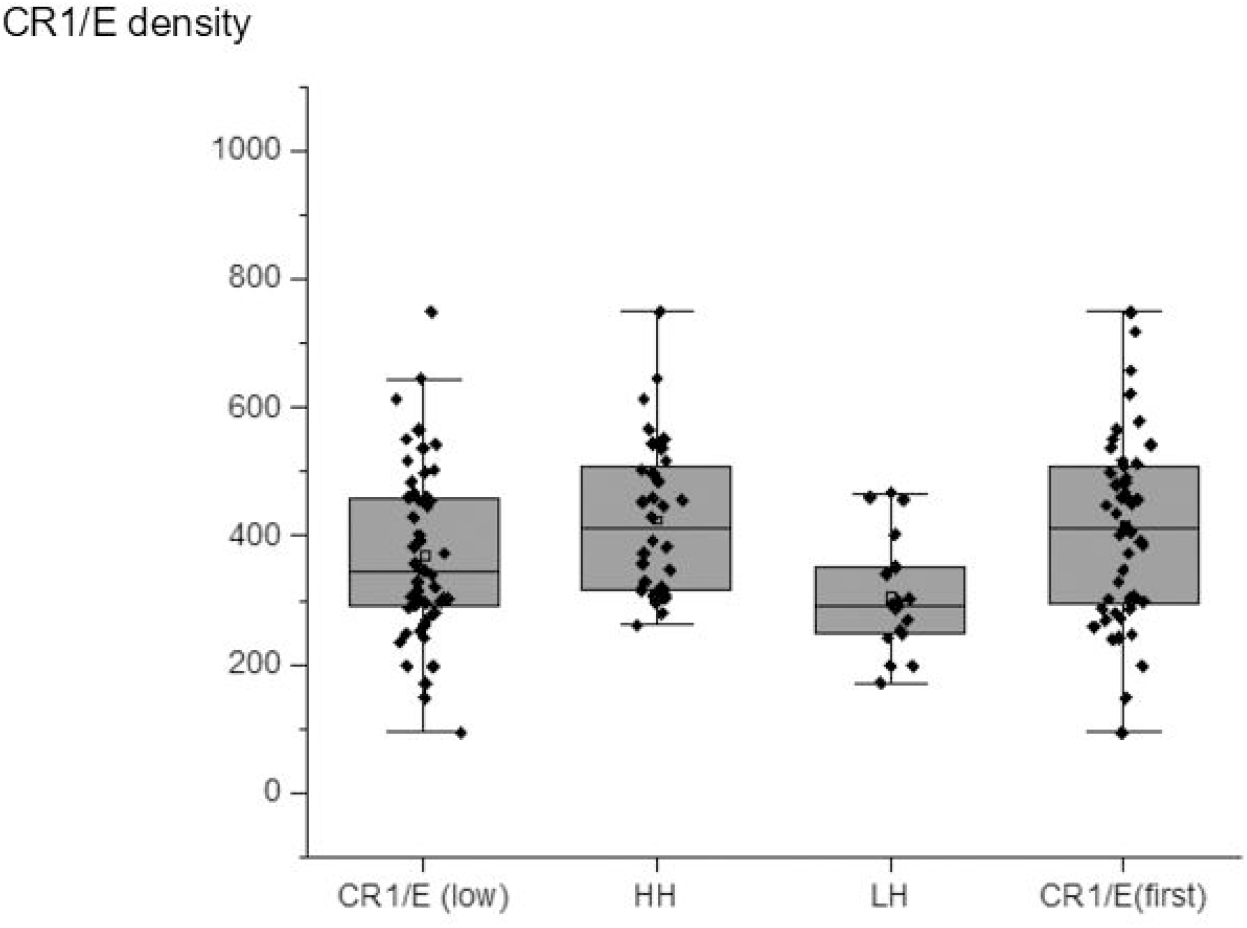
Comparison of CR1/E density among COVID-19 patients according to CR1 density genotypes. CR1/E (low): from patients at the lowest value during longitudinal follow-up. (Mean = 372 SD = 134 N = 52). CR1/E (first): CR1 on erythrocyte from patients at the first determination (Mean = 418 SD = 162 N = 52). Among CR1/E (low) HH: patients homozygous for the high density genotype (Mean = 426 SD = 122 N = 32). LH patients heterozygous for the high and low density genotypes (Mean = 308 SD = 93 N = 17). Means are depicted as open squares. The 3 patients homozygous for the LL low density genotype are not shown. Heterozygous patients displayed lower CR1 density values than homozygous patients. Wilcoxon median test (p<0,001).

**Fig. 3.**
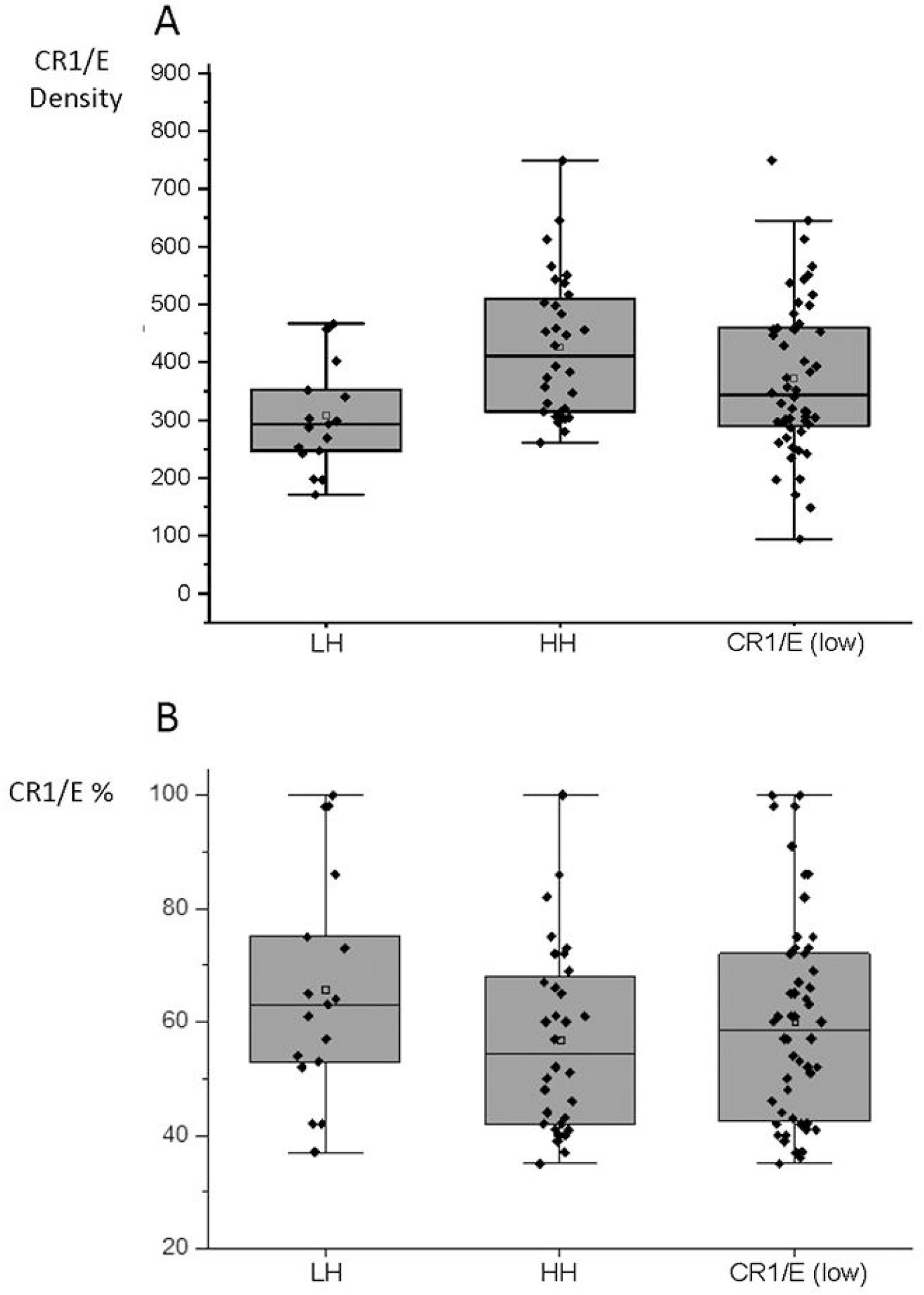
Comparison of the decrease of CR1/E density at first determination among COVID-19 patients according to CR1 density genotypes. (**A)** Among CR1/E (low) from patients at the lowest value during longitudinal follow-up (Mean = 372 SD = 134 N = 52). HH: patients homozygous for the high density genotype (Mean = 426 SD = 122 N = 32). LH patients heterozygous for the high and low density genotypes (Mean = 308 SD = 93 N = 17). (**B)** CR1/E%: CR1/E were expressed as the percentage of the mean CR1/E density predicted from the genotype of a given individual. Among CR1/E (low) from patients at the lowest value during longitudinal follow-up (Mean = 60 SD = 18.3 N = 52). HH% (Mean = 57 SD = 16.3 N = 32); LH% (Mean = 66 SD = 19.9 N = 17). Means are depicted as open squares. Proportionally, patients with the HH high density genotype displayed a more important loss of CR1/E than LH heterozygous patients. Wilcoxon median test (p< 0.001).

### 3.2 Decrease of CR1/E density among COVID-19 patients is independent of the inherited genetic control by HindIII density polymorphism rs11118133

During the course of the disease, CR1/E continued to decrease (CR1/E (Low)) when compared to initial CR1/E determination (CR1/E (First)) (Fig. 2, supplemental Fig. 1).

The *Hin*dIII polymorphism rs11118133 was determined for every patient. As expected, the three genotypes HH, HL, or LL were similarly represented in 52 patients and 87 elderly controls (HH = 62.7%, HL = 31.4%, LL = 5.9% and HH = 63.2%, HL = 32.2%, LL= 4.6% respectively). H standing for high, L low and HL intermediate levels.

The allele frequency was also the same in COVID-19 patients (H = 78.73%; L = 21.57%) and in healthy individuals is (H = 79.31%; L = 20.69%). No inherited CR1 susceptibility factor for COVID-19 was found.

As expected CR1/E values from “high” (HH genotype) were higher than CR1/E from “medium” (HL genotype) (Fig. 2, Fig. 3A).

Expression of the CR1/E density as CR1/E% of the expected density for a given individual allowed us to observe that both HH homozygous individuals for the rs11118133 density polymorphism “high”, or LH or “medium”, inherited CR1/E density groups exhibited a decrease of CR1/E. Although the “medium” group exhibited less CR1/E than the “high” group, conversion to CR1/E% showed that the “high” group exhibited a larger decrease in CR1/E% (HH = 57% CR1/E% versus LH = 66% CR1/E% (Fig. 3B). This is in accordance with the catabolic rate of CR1/E *in vivo*, which is higher on E displaying more CR1 that on those of lower CR1 density (Dervillez et al., 1997). This finding may be explained by established dynamics of increased catabolic rate of clearance of CR1 on erythrocytes, or reticulocytes *in vivo* (Cohen et al., 1992).

### 3.3 Decrease of CR1/E density according to the age of patients

Among all patients, age was a major predictive factor of decreased CR1/E. We used 350 CR1/E as a low-level cut-off of CR1/E. We looked at levels of CR1/E in HH or HL bearing genotype patients and there was a significant difference of 7.7 years of age between those with high versus low values (Mean age = 71.24, Days = 17966 SD = 2319 N = 20 versus Mean age = 63.53, Days = 20778 SD = 5206 = 32. Student’s t-test p < 0,015) Medians were also different by 6.19 Year (66.40 versus 70.59) (Fig. 4). That can also be visualized by plotting CR1/E versus dates of birth. (supplemental Fig. 4).

**Fig. 4.**
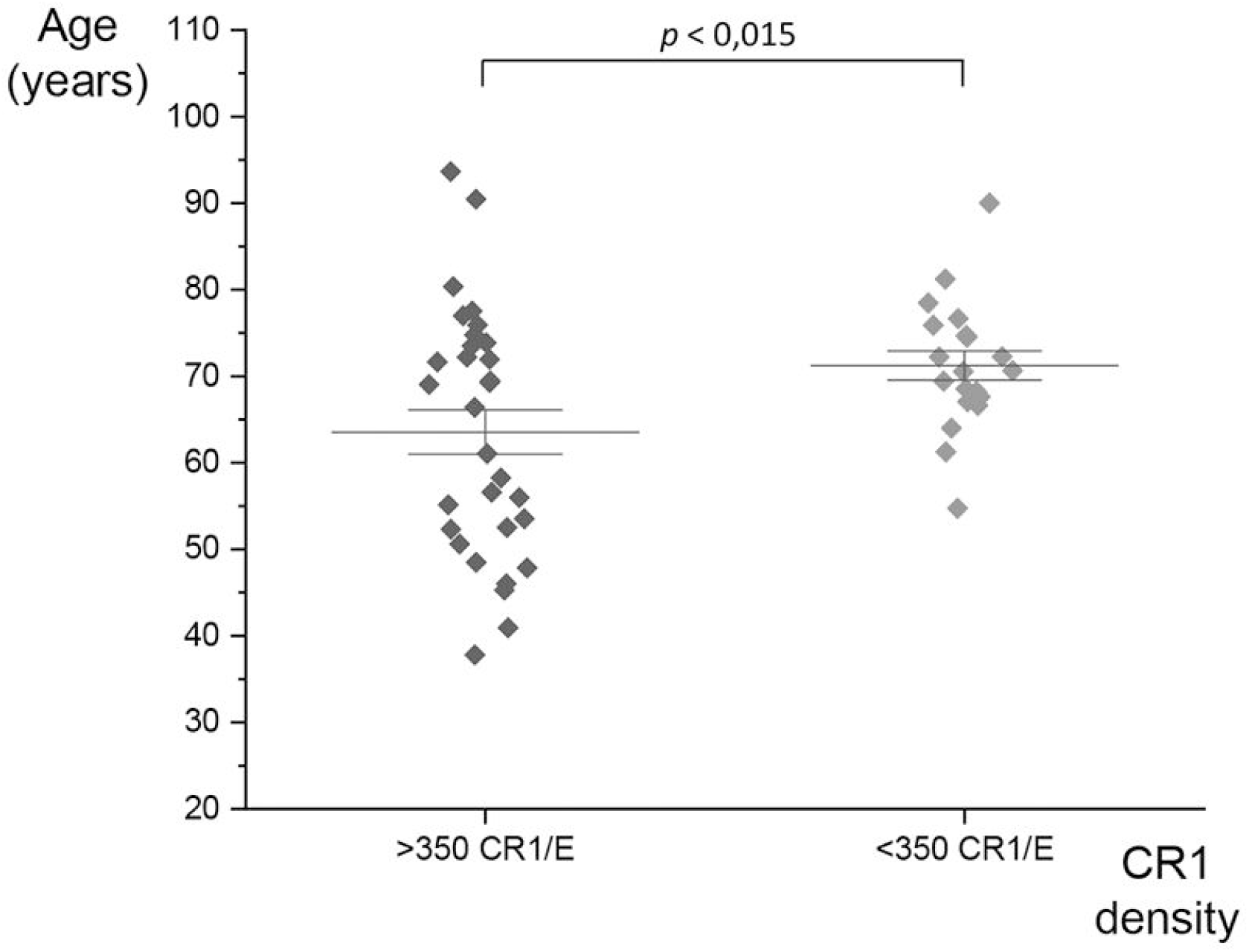
Comparison of age of COVID-19 patients and CR1/E. At a cut-off of density greater than 350 (>350 CR1/E) CR1/E antigenic sites or below 350 (<350 CR1 / E). The standard error of the mean is also represented. A significant difference of 7.7 years is observed when compared COVID-19 patients > 350 CR1 / E (Mean age = 63.53, Days = 20778 SD = 5206 N = 32) to COVID-19 patients < 350 CR1 / E (Mean age = 71.24, Days = 17966 SD = 2319 N = 20). Student’s t-test p < 0,015).

### 3.4 Large C4d and fewer C3b/C3bi deposits are observed in COVID-19 patients

C3b /C3bi and C4d deposits were detected at E surface of 114 samples from 52 COVID-19 patients. In most patients, the levels observed were greater than the basal levels observed in healthy subjects. Thus, 25% of COVID-19 patients had deposits of C3b / C3bi and 83% of COVID-19 patients had huge deposits of C4d on the surface of their E.These large deposits were in most patients several fold more abundant than C4d deposits found in normal individuals, p<10^−6^ (Fig.5). The values for the control, healthy population were converted to the scale of the C4d deposit measurement used in the present study, using the values of 2 healthy individuals tested in both studies: Mean = 499 SD = 59 N = 88 (Fig. 5B). There is a trend towards a correlation between the deposition of C4d and the low density of CR1/E. (supplemental Fig. 6).

**Fig. 5.**
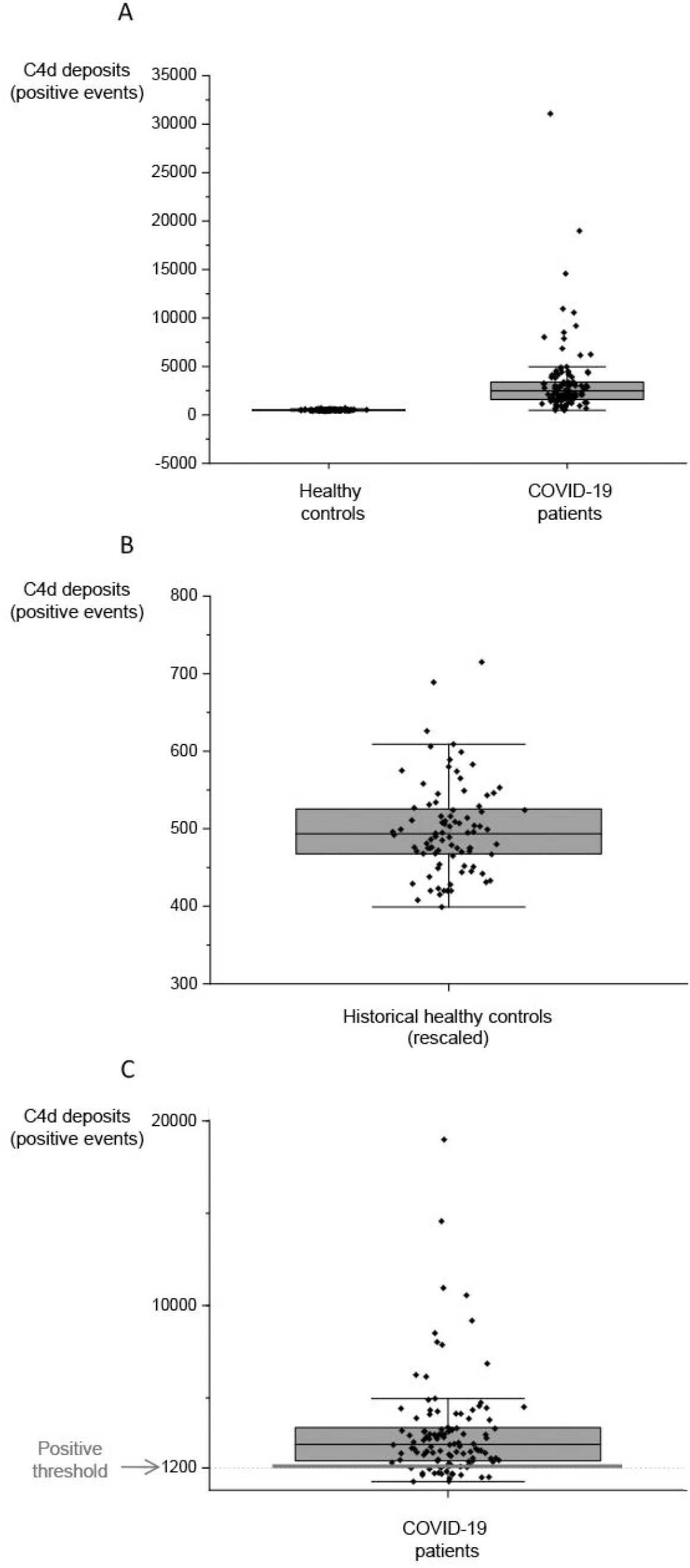
C4d deposits on erythrocyte from COVID-19 patients and healthy individuals. **(A)** Distribution of C4d staining on erythrocyte from COVID**-**19 patients and historical healthy controls (Mean = 499 SD = 58.92 N = 88) rescaled to the scale of the present study using values of 2 patients tested in both studies. Y axis: Positive events. (**B**) Levels of C4d staining on erythrocyte in healthy individuals. The scale has been adapted to expand the limited range of C4d staining in normal individuals. (**C)** Levels of C4d staining on erythrocyte in patients. The threshold of positivity used is depicted in grey.

**Fig. 6.**
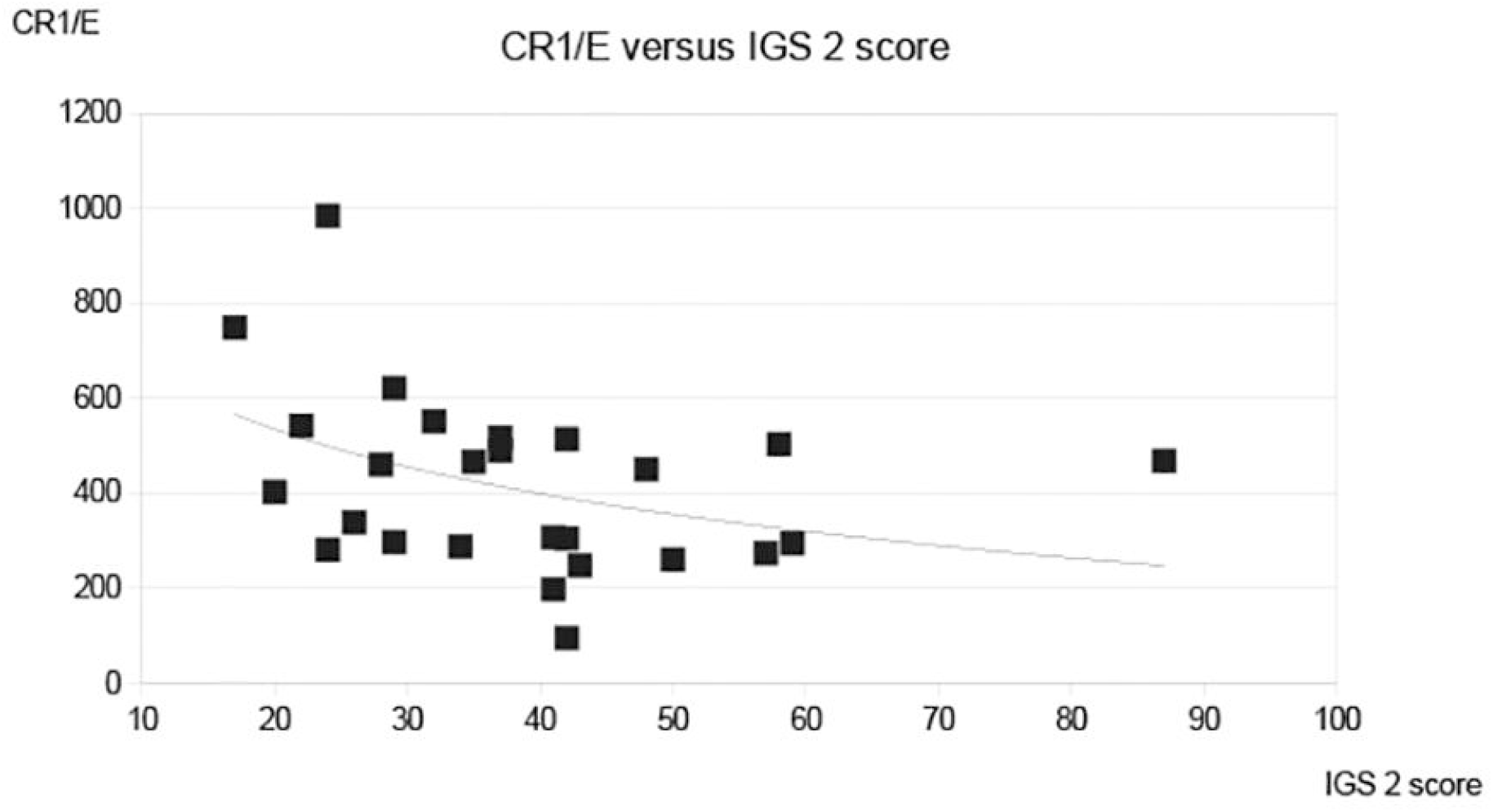
Comparison of CR1/E density at first determination to acute physiological score IGS2. CR1/E density is inversely related to IGS 2 severity index, N=27 f(x) = -196 ln (x) +1120 R^2^= 0.16.

No concomitant crease in C3b /C3bi deposits was found on the E samples with the highest C4d deposits.

### 3.5 Acquired decrease in CR1/E among patients is related to clinical severity of the disease

There are several observations that indicate that the magnitude of the acquired decrease of CR1/E that we observed is likely to be associated with more severe disease and possibly mortality. Deceased patients exhibited a trend toward lower CR1/E levels (Fig. 1), CR1/E was lower in more aged patients known to be at higher risk for more severe disease (supplemental Fig. 4 and supplemental Fig. 5), the physiological index IGS2, (Le Gall J. et al., 1983) at ICU admission was inversely related to the CR1/E level (Fig. 6), and CR1/E was lower among patients with refractory hypoxemia (Fig. 7)

**Fig. 7.**
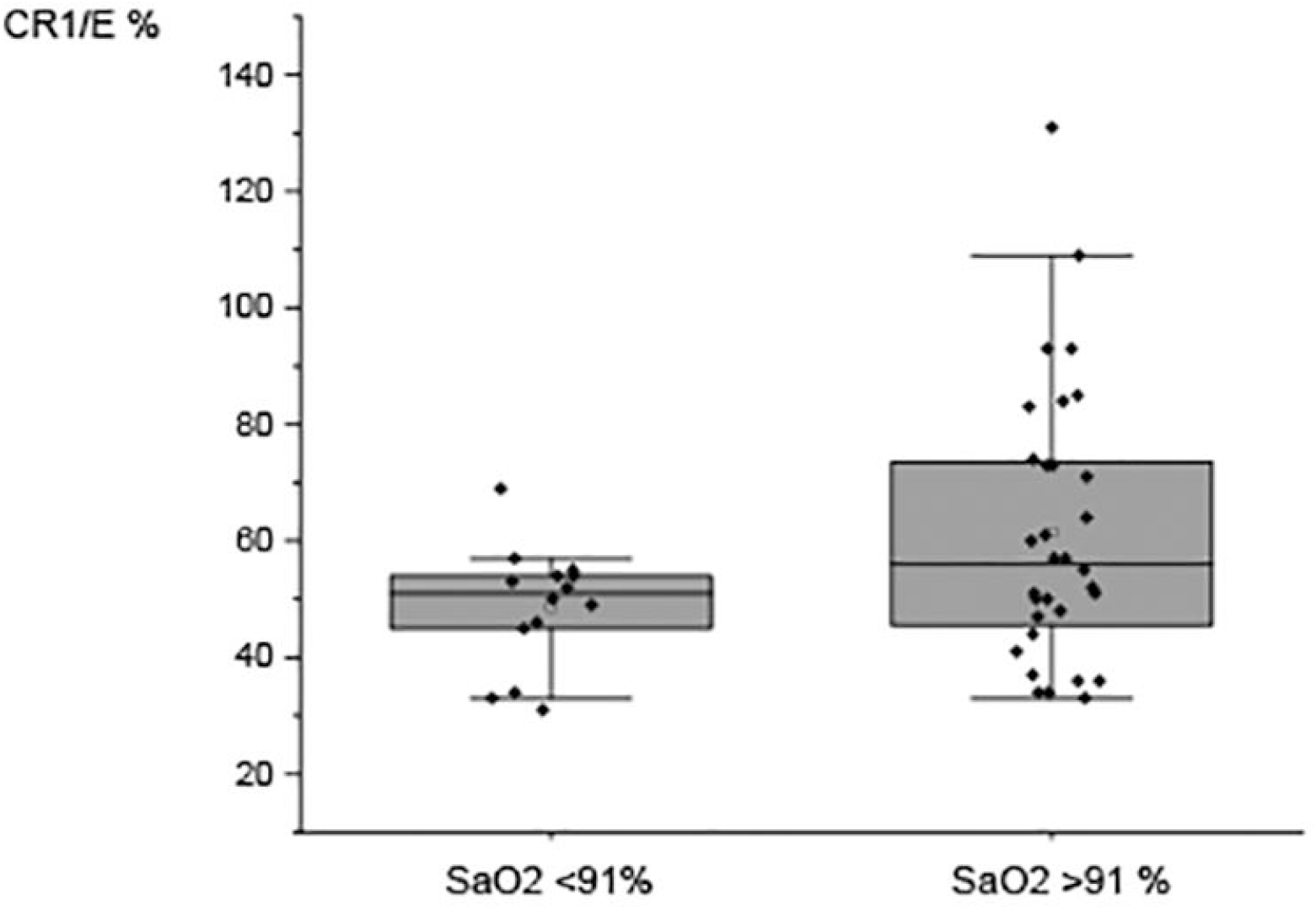
Comparison of CR1/E% and SaO2 between COVID-19 patients. SaO2 <91% (Mean = 48.71 SD = 10.39 N = 14) versus SaO2 greater than or equal to 91% (Mean = 61.47 SD = 23.50 N = 32). Means are depicted as open squares. Patients with SaO2 lower than 91% under the best O_2_ therapy were considered as refractory hypoxemia. A reduced erythrocyte CR1 density was observed in severely affected patients with SaO2 <91%. Wilcoxon signed rank test p<0.01.

## 4. Discussion

As we expected from what is known about the clinical course of severe COVID-19, and the role of complement activation in the pathophysiology of the severe COVID-19, we hypothesized that the regulation of complement activation on E was involved and we found evidence that it does. This is in keeping with the role of E complement receptors, mostly CR1, as the main “cooling system” of complement activation and inflammation on endothelial cells in capillaries,

A deterioration in the clinical course of the disease after the 7th day of symptoms has been described in COVID-19 patients (Hozhabri et al., 2020). We considered that the complement system, by activating during the first days of the SARS-CoV-2 infection, during the innate immune response, could be modified by the virus, causing its deregulation, leading to increased activation of specific, critical components of the immune system and deterioration of the patient’s state of health during a second phase.

We assessed complement activation by analyzing the presence of C3b/ C3bi and C4d residual molecules of complement activation on the surface of patients’ E. We focused in the CR1 receptor on E, which plays a key role in inhibiting complement activation and in the clearance of immune complexes from the blood.

We conducted this study in the context of the French lock-down with the practical limitations of a limited number of patients with variable follow-up. These results were obtained from patients at an advanced stage of the disease, and were not from a longitudinal study from the early onset of the disease that would have been more informative, but impracticable, in the context of the epidemic. Correlations to other biological or clinical parameters that can only be considered as trends that would need to be confirmed in further studies have been given in the section supplemental materials.

Most of the patients only had deposits of C4d on the E. This agrees with the work of A. Berzuini *et al*. who detected C3 in 12% only of their patients, using anti-C3d, to test COVID-19 patients who were positive on the direct antiglobulin test (DAT), consistent with our observation that the C4d deposits we found in many cases were isolated, without concomitant C3 deposits (Berzuini et al., 2020). It also coincides with the results observed in kidney transplant rejection (Golocheikine et al., 2010; Haidar et al., 2012), and clinical flare-ups of SLE (Manzi et al., 2004). Peri-vascular C4d deposits in chronic vascular rejection are also observed without C3 or Ig deposits in most cases.

The deposits of C4d on the E of patients with COVID-19 might reflect a phenomenon in the peripheral blood that is also occurring in capillaries, resulting in end-organ damage seen clinically. Recently, Holter *et al*. from Norway reported on complement activation in the plasma fluid phase of COVID-19 patients. They found a trend of elevated plasma C4d levels in COVID-19 patients with hypoxemia (Holter et al., 2020). This trend of increased C4d was in the same direction we found in our subjects with hypoxemia. Taken together with our data, there are two independent cohorts using different approaches, Holter *et al*. looking at the fluid phase as an instantaneous measurement of C4d or in our study on E as a cumulative parameter, that reflects long-terms changes during the diseases. Measuring complement fragments in the serum gives a snapshot at a given time, whereas measuring CR1/E gives a picture of the patient’s response to infection over a longer duration, like HbA1c in diabetes mellitus.

C3b /C3bi and C4d have been detected by a flow cytometry using sensitive methods that allow evaluation of the low levels found in normal individuals. C3b /C3bi or C4d staining patterns in patients were quite different. C3b /C3bi deposits remained in the normal range in ¾ of patients, ¼ exhibited larger deposits.

The number of C4d positive E cells was above values observed in normal individuals in 83% of the tested samples. No increase in C3b /C3bi deposits were found on the E samples with the highest C4d deposits. Taken together, these data suggest that C4 activation is independent from the C3, suggesting that it was not activation by the C3 loop from the alternative pathway.

We also saw a decrease in the number of CR1 receptors on the surface of the E of COVID-19 patients. The allele frequencies of the rs11118133 SNP associated with the CR1/E density polymorphism were not different from those of the general population. Therefore, this decrease in CR1 receptors was not due to genetic factors in our cohort. However, the acquired decrease of CR1/E observed in patients was proportionally larger among individuals with the HH polymorphism that is associated with a higher inherited CR1/E density. This is in accordance with the larger decrease of CR1 during E life among individuals expressing a high density of CR1/E, as well as among patients with Alzheimer’s disease who had the high-density genotype (Mahmoudi et al., 2018).

With less CR1, complement inhibition is reduced, which enables increased, ongoing activation of the complement cascade, amplifying the immune response. In addition, with lower levels of CR1, immune complexes are not cleared as efficiently from the blood and so will accumulate and participate in tissue inflammation, which could explain the phenomena of thrombosis seen in COVID-19 patients (Oikonomopoulou et al., 2012).

The magnitude of the acquired decrease of CR1/E appeared to be associated with increased severity of the disease. Deceased patients exhibited a trend toward lower CR1/E levels, CR1/E was lower in older patients known to be at risk for more severe disease, the physiological index IGS2 at ICU admission was inversely related to CR1/E level, D-Dimer levels showed a trend in the same direction, and CR1/E was lower among patients with a refractory hypoxemia.

The decreased CR1/E among elderly patients was strikingly similar to the now well-known effect of age on the mortality curve of COVID-19 patients (Williamson et al., 2020). (supplemental Fig. 5). The hypothesis can be made that some as yet unknown factor changes with age that determines the magnitude of the deleterious inflammation and complement activation in COVID-19 patients.

The decreased CR1/E density presented here, the detection of virus spikes, and the presence of C3 (Metthew Lam et al., 2020) or C4 fragments on E among COVID-19 patients, are likely to be different aspects of the same phenomenon.

From these data we make the hypothesis that in COVID-19 patients, CR1 on E captures complement fragment coated-virus, virus-containing immune complexes or cellular debris, as well as inactivates the C3 loop complement amplification on all of them. When overwhelmed by the amount of immune complexes they are required to handle and by the magnitude of complement activation at E surface, E are shedding CR1 through exocytosis or proteolysis, resulting in a decrease of CR1 density on E. Complement fragment deposits, mostly C4d, then accumulate more on less efficiently defended E. This leads to a cycle that amplifies the immune response to the virus in a vicious cycle.

Our results showing an acquired decrease in CR1/E and the presence of C4d/E deposits in most patients confirms the role of complement in COVID-19. C4d/E deposition, which was found to be elevated in most ICU COVID-19 patients, might be an early signal of vascular damage, a surrogate marker for an increased risk of end organ damage and thrombosis, indicating the potential use of these parameters for patient monitoring. CR1/E assessment, when coupled to rs11118133 SNP assessment and C4d/E deposits could be also evaluated as a predictive factor in COVID-19 patients. These results also suggest that complement regulatory molecules could be useful in the treatment of COVID-19. This may include the use of CR1, or CR1-like molecules with the aim of down regulating complement activation and inflammation for therapy, either in the liquid phase or increasing the CR1 density on E to the higher range observed in other primates (Oudin et al., 2000).

## Data Availability

We can provide upon request from researchers fresh reference E without charge for the calibration curve of Flow Cytometry measurement of CR1/E. As well as assistance for setting-up methologies used from the corresponding Author.

## Abbreviations

CR1: complement receptor type 1
CR1/E: complement receptor type 1 per erythrocyte
CR1/E%: complement receptor type 1 per erythrocyte as percentage of the CR1/E density expected from the CR1 density polymorphism genotype of the patient
C3: complement fragment 3
C4: complement fragment 4
C1q: complement component 1q
C1r: complement component 1r
C1s: complement component 1s
C3a: complement component 3a
C3b: complement component 3b
C4a: complement component 4a
C4d: complement component 4d
C5a: complement component 5a
DAT: direct antiglobulin test
E: erythrocytes
FITC: fluorescein iso-thio-cyanate
HbA_1c_: glycosylated hemoglobin type A
ICU: intensive care unit
MBL: mannan binding lectin
MASP1: mannan binding lectin serine protease1 PE phyco-erythrin
Sa02: oxygen saturation
SLE: systemic lupus erythematosus

## Acknowledgments

Ms B. Reveil for her highly appreciated technical and moral assistance. Dr R. Levinson for helpful advice.

This work was partly funded by the “Association pour le Développement de la Microbiologie et de l’Immunologie Ré-moises” (ADMIR) non-profit Organization.

## Authorship

Contribution: A. K.: Flow Cytometry and biological data processing, N. S.: Clinical data processing, S. A.: Flow Cytometry, T. T.: Logistic and data processing, A. G. and J. C.: Care of ICU patients and clinical data processing, R. M. : Management of the cohort of aged individuals, F. B.-S.: Care of non ICU patients, L. K. and D. J.: Management of the biological bank and data processing, J.H.M. C.: Conception of the study and synthesis of results

## Footnotes

The Rheims University Hospital has obtained the authorization to create plasma and serum libraries from the Human Protection Committee (CPP Est III, national number: 2020-A01093-36) in order to study the soluble Vascular Endothelial Growth Factor (sFlt-1) receptor, in intensive care patients with severe COVID-19 pneumonitis. The patients or their families signed an informed consent form to participate, specifying that other elements, such as the red blood cells that were to be thrown away, could be used for research for other purposes, in the context of the COVID-19 epidemic. Blood donors were healthy unpaid Volunteers who signed the general informed consent from the national Établissement Français du Sang (EFS). Aged patients were from a cohort with individual informed consents approved by the regional ethics committee (CPP Est II), under the protocol number 2011-A00594-37. Consents of the healthy volunteers of the C4d/E reference cohort were also approved by the local ethics committee in 2006.

## References

Atkinson J.P., 1988. Origin of the Fourth Component of Complement Related Chido and Rodgers Blood Group Antigens. Complement. 5, 65–76.

Berzuini, A., Bianco, C., Paccapelo, C., Bertolini, F., Gregato, G., Cattaneo, A., et al., 2020. Red cell–bound antibodies and transfusion requirements in hospitalized patients with COVID-19. Blood 136, 766–768. https://doi.org/10.1182/blood.2020006695

Birmingham, D. J., Chen, W., Liang, G., Schmitt, H. C., Gavit, K., Nagaraja, H. N., 2003. A CR1 polymorphism associated with constitutive erythrocyte CR1 levels affects binding to C4b but not C3b. Immunology. 108, 531–538.

Carvelli J., Demaria O., Vély F., Batista L., Benmansour N.C., Fares J., et al., 2020. Association of COVID-19 inflammation with activation of the C5a-C5aR1 axis. Nature. doi: 10.1038/s41586-020-2600-6.

Cavaillon J.M., Sansonetti P., Goldman M., 2019. 100th Anniversary of Jules Bordet’s Nobel Prize: Tribute to a Founding Father of Immunology. Front Immunol. 10: 2114. doi: 10.3389/fimmu.2019.02114.

Cohen, J.H., Aubry, J.P., Jouvin, M.H., Wijdenes, J., Bancherau, J., Kazatchkine, M., Revillard, J.P., 1987. Enumeration of CR1 complement receptors on erythrocytes using a new method for detecting low density cell surface antigens by flow cytometry. J. Immunol. Methods. 99, 53–58.

Cohen, J.H., Lutz, H.U., Pennaforte, J.L., Bouchard, A., Kazatchkine, M.D., 1992. Peripheral catabolism of CR1 (the C3b receptor, CD35) on erythrocytes from healthy individuals and patients with systemic lupus erythematosus (SLE). Clin. Exp. Immunol. 87, 422–428.

Cornacoff, J.B., Hebert, L.A., Smead, W.L., VanAman, M.E., Birmingham, D.J., Waxman, F.J., 1983. Primate erythrocyte-immune complex-clearing mechanism. J. Clin. Investig. 71, 236–247.

Cornillet P., Philbert F., Kazatchkine M.D., Cohen J.H., 1991. Genomic determination of the CR1 (CD35) density polymorphism on erythrocytes using polymerase chain reaction amplification and HindIII restriction enzyme digestion. J. Immunol. Methods. 136, 193–197.

Dervillez, X., Oudin, S., Libyh, M.T., Reveil, B., Philbert, F., Bougy, F., Pluot, M., Cohen, J.H., 1997. Catabolism of the human erythrocyte C3b /C4b receptor (CR1, CD35): vesiculation and/or proteoly-sis? Immunopharmacology 38, 129–140.

Dobson, N. J., Lambris, J. D., Ross, G. D., 1981. Characteristics of isolated erythrocyte complement receptor type one (CR1, C4b-C3b receptor) and CR1-specific antibodies. J. Immunol. 126, 693–698.

Dykman, T. R., Hatch, J. A., Aqua, M. S., Atkinson, J. P., 1985. Polymorphism of the C3b/C4b receptor (CR1): characterization of a fourth allele. J. Immunol. 134, 1787–1789.

Edberg J.C., Wright E., Taylor R.P., 1987. Quantitative analyses of the binding of soluble complement-fixing antibody/dsDNA immune complexes to CR1 on human red blood colis. J. Immunol. 139, 3739–3747.

Fearon, D. T., 1979. Regulation of the amplification C3 convertase of human complement by an inhibitory protein isolated from human erythrocyte membrane. Proc. Natl. Acad. Sci. USA. 76, 5867–5871.

Fearon, D. T., 1980. Identification of the membrane glycoprotein that is the C3b receptor of the human erythrocyte, polymorphonuclear leukocyte, B lymphocyte, and monocyte. J. Exp. Med. 152, 20–30.

Ferreira A., 1980. The murine H-2.7 specificity is an antigenic determinant of C4d, a fragment of the fourth component of the complement system. J. Exp. Med. 151, 1424–1435.

Feucht, H.E., Felber, E., Gokel, M.J., Hillebrand, G., Nattermann, U., Brockmeyer, C., et al., 1991. Vascular deposition of complement-split products in kidney allografts with cell-mediated rejection. Clin. Exp. Immunol. 86, 464–470.

Freysdottir J., Sigfusson A., 1991. A flow cytometric assay for measuring complement receptor 1 (CR1) and the complement fragments C3d and C4d on erythrocytes. J. Immunol. Methods 142, 45–52.

Gasque, P., Chan, P., Mauger, C., Schouft, M. T., Singhrao, S., Dierich, M. P. et al., 1996. Identification and characterization of complement C3 receptors on human astrocytes. J. Immunol. 156, 2247–2255.

Ghiran, I., Barbashov, S. F., Klickstein, L. B., Tas, S. W., Jensenius, J. C., Nicholson-Weller, A., 2000. Complement receptor 1/CD35 is a receptor for mannan-binding lectin. J. Exp. Med. 192, 1797–1808.

Giles C.M., Robson T., 1991. Immunoblotting human C4 bound to human erythrocytes in vivo and in vitro. Clin. Exp. Immunol., 84, 263–269.

Golocheikine, A., Nath, D.S., Basha, H.I., Saini, D., Phelan, D., Aloush, A. et al., 2010. Increased erythrocyte C4D is associated with known alloantibody and autoantibody markers of antibody-mediated rejection in human lung transplant recipients. J. Heart Lung Transplant. 29, 410–416.

Gralinski, L.E., Sheahan, T.P., Morrison, T.E., Menachery, V.D., Jensen, K., Leist, S.R., Whitmore, A., Heise, M.T., Baric, R.S., 2018. Complement Activation Contributes to Severe Acute Respiratory Syndrome Coronavirus Pathogenesis. Mbio 9, e01753–18.

Haidar, F., Kisserli, A., Tabary, T., McGregor, B., Noel, L.H., Réveil, B., et al., 2012. Comparison of C4d Detection on Erythrocytes and PTC-C4d to Histological Signs of Antibody-Mediated Rejection in Kidney Transplantation. Am. J. Transplant. 12, 1564–1575.

Herrera, A.H., Xiang, L., Martin, S.G., Lewis, J., Wilson, J.G., 1998. Analysis of complement receptor type 1 (CR1) expression on erythrocytes and of CR1 allelic markers in caucasian and african american populations. Clin. Immunol. Immunopathol. 87, 176–183.

Holter JC, Pischke SE, de Boer E, Lind A, Jenum S, Holten AR, et al., 2020. Systemic complement activation is associated with respiratory failure in COVID-19 hospitalized patients. Proc. Natl. Acad. Sci. U S A. 117, 25018–25025. doi: 10.1073/pnas.2010540117.

Horgan, C., Taylor, R. P., 1984. Studies on the kinetics of binding of complement-fixing dsD-NA/anti-dsDNA immune complexes to the red blood cells of normal individuals and patients with systemic lupus erythematosus. Arthritis & Rheumatology. 27, 320–329.

Hozhabri H., Piceci Sparascio F., Sohrabi H., Mousavifar L., Roy R., Scribano D., et al., 2020. The Global Emergency of Novel Coronavirus (SARS-CoV-2): An Update of the Current Status and Fore-casting. Int. J. Environ. Res. Public Health. 17(16): 5648. doi: 10.3390/ijerph17165648

Iida, K., Nussenzweig, V., 1981. Complement receptor is an inhibitor of the complement cascade. J. Exp. Med. 153, 1138–1150.

Jouvin, M.H., Rozenbaum, W., Russo, R., Kazatchkine, M.D., 1987. Decreased expression of the C3b /C4b complement receptor (CR1) in AIDS and AIDS-related syndromes correlates with clinical subpopulations of patients with HIV infection. AIDS 1, 89–94.

Kerepesi L.A., Hess J.A., Nolan T.J., Schad G.A., Abraham D., 2006. Complement component C3 is required for protective innate and adaptive immunity to larval strongyloides stercoralis in mice. J. Immunol. 176, 4315–4322.

Kerr M.A., 1981. The complement system. Biochem. Educ. 9, 82–88.

Kerr Anderson, W.I., Stroud, R.M., 1979. Generation and enrichment of C4d in whole serum. J. Immunol. Methods. 29, 323–330.

Kisserli, A., Audonnet, S., Duret, V., Tabary, T., Cohen, J.H.M., Mahmoudi, R., 2020. Measuring Erythrocyte Complement Receptor 1 Using Flow Cytometry. JoVE. https://doi.org/10.3791/60810

Klickstein, L. B., Barbashov, S. F., Liu, T., Jack, R. M., Nicholson-Weller, A., 1997. Complement receptor type 1 (CR1, CD35) is a receptor for C1q. Immunity. 7, 345–355.

Leddy J.P., Frank M.M., Gaither T., Baum J. and Klemperer M.R., 1974. Hereditary deficiency of the sixth component of complement in man. I. Immunochemical, biologic, and family studies. J. Clin. Invest. 53, 544–553.

Lee, K.C., Chang, C.Y., Chuang, Y.C., Sue S.H., Chu, T.W., Chen, R. J., et al., 2008. Measurement of human erythrocyte C4d to erythrocyte complement receptor 1 ratio in cardiac transplant recipients with acute symptomatic allograft failure. Transplant Proc. 40, 2638–2642.

Le Gall J., Loirat P., Alperovitch A., 1983. Simplified Acute Physiological Score for intensive care patients. Lancet 2, 741).

Magro, C., Justin-Mulvey, J., Berlin, D., Nuovo, G., Salvatore, S., Harp, J., et al., 2020. Complement Associated Microvascular Injury and Thrombosis in the Pathogenesis of Severe COVID-19 Infection: A Report of Five Cases. Transl. Res. 220, 1–13.

Mahmoudi, R., Kisserli, A., Novella, J.L., Donvito, B., Dramé, M., Réveil, B. et al., 2015. Alzheimer’s disease is associated with low density of the long CR1 isoform. Neurobiol. Aging. 36, 1766.e5-1766.e12.

Mahmoudi, R., Feldman, S., Kisserli, A., Duret, V., Tabary, T., Bertholon, L.A et al., 2018. Inherited and Acquired Decrease in Complement Receptor 1 (CR1) Density on Red Blood Cells Associated with High Levels of Soluble CR1 in Alzheimer’s Disease. Int. J. Mol. Sci. 19, pii: E2175.

Manzi S., Navratil J.S., Ruffing M.J., Liu C.C., Danchenko N., Nilson S.E. et al., 2004. Measurement of erythrocyte C4d and complement receptor 1 in systemic lupus erythematosus. Arthritis. Rheum. 50, 3596–3604.

Metthew Lam, L., Murphy, S.J., Kuri-Cervantes, L., Weisman, A.R., Ittner, C.A.G., Reilly, J.P., et al., 2020. Erythrocytes Reveal Complement Activation in Patients with COVID-19. https://doi.org/10.1101/2020.05.20.20104398.

Moulds, J. M., Moulds, J. J., Brown, M., Atkinson, J. P., 1992. Antiglobulin testing for CR1-related (Knops/McCoy/Swain-Langley/York) blood group antigens: negative and weak reactions are caused by variable expression of CR1. Vox Sanguinis. 62, 230–235.

Murakami Y., Imamichi T., Nagasawa S., 1993. Characterization of C3a anaphylatoxin receptor on guinea-pig macrophages. Immunology. 79, 633–638.

Oikonomopoulou K., Ricklin D., Ward P.A., and Lambris J.D., 2012. Interactions between coagulation and complement—their role in inflammation. Semin. Immunopathol. 34, 151–165.

Oudin, S., Libyh, M.T., Goossens, D., Dervillez, X., Philbert, F., Réveil, B., Bougy, F., Tabary, T., Rouger, P., Klatzmann, D., Cohen, J. H., 2000. A soluble recombinant multimeric anti-Rh (D) single-chain Fv/CR1 molecule restores the immune complex binding ability of CR1-deficient erythrocytes. J. Immunol. 164, 1505–1513.

Pascual, M., Steiger, G., Sadallah, S., Paccaud, J. P., Carpentier, J. L., James R. et al., 1994. Identification of membrane-bound CR1 (CD35) in human urine: evidence for its release by glomerular po-docytes. J. Exp. Med. 179, 889–899.

Pham, B. N., Kisserli, A., Donvito, B., Duret, V., Reveil, B., Tabary, T et al., 2010. Analysis of complement receptor type 1 expression on red blood cells in negative phenotypes of the Knops blood group system, according to CR1 gene allotype polymorphisms. Transfusion. 50, 1435–1443.

Reynes, M., Aubert, J. P., Cohen, J. H., Audouin, J., Tricottet, V., Diebold, J. et al., 1985. Human follicular dendritic cells express CR1, CR2, and CR3 complement receptor antigens. J. Immunol. 135, 2687–2694.

Risitano, A.M., Mastellos, D.C., Huber-Lang, M., Yancopoulou, D., Garlanda, C., Ciceri, F., Lambris, J.D., 2020. Complement as a Target in COVID-19? Nat. Rev. Immunol. 20, 343–344.

Rodriguez de Cordoba, S., Rubinstein, P., 1986. Quantitative variations of the C3b/C4b receptor (CR1) in human erythrocytes are controlled by genes within the regulator of complement activation (RCA) gene cluster. J. Exp. Med. 164, 1274–1283.

Ross, G. D., Winchester, R. J., Rabellino, E. M., Hoffman, T., 1978. Surface markers of complement receptor lymphocytes. J. Clin. Investig. 62, 1086–1092.

Ross, G. D., Newman, S. L., Lambris, J. D., Devery-Pocius, J. E., Cain, J. A., Lachmann, P. J., 1983. Generation of three different fragments of bound C3 with purified factor I or serum. II. Location of binding sites in the C3 fragments for factors B and H, complement receptors, and bovine conglutinin. J. Exp. Med. 158, 334–352.

Sacks S.H., Chowdhury P., Zhou W., 2003. Role of the complement system in rejection. Curr. Opin. Immunol. 15, 487–492.

Schreiber, R. D., Pangburn, M. K, Muller-Eberhard, H. J., 1981. C3 modified at the thiolester site: acquisition of reactivity with cellular C3b receptors. Bioscience Reports. 1, 873–880.

Tilley, C.A., Romans, D.G., Crookston, M.C., 1978. Localisation of Chido and Rodgers determinants to the C4d fragment of Human C4. Nature 276, 713–715.

Van Del Elsen J.M., Martin A., Wong V., Clemenza L., Rose D.R. et al., 2002. X ray crystal structure of the C4d fragment of human complement component C4, J. Mol. Biol. 322, 1103–1115.

Waitumbi, J.N., Donvito, B., Kisserli, A., Cohen, J.H.M., Stoute, J.A., 2004. Age-related changes in red blood cell complement regulatory proteins and susceptibility to severe malaria. J Infect Dis. 190, 1183–1191.

Walport M.J., 2001. Complement. Second of two parts. N. Engl. J. Med. 344, 1140–1144.

Walport M.J., 2001. Complement. First of two parts. N. Engl. J. Med. 344, 1058–1066.

Wang, F.S., Chu, F.L., Jin, L., Li, Y.G., Zhang, Z., Xu, D., Shi, M., Wu, H., Moulds, J.M., 2005. Acquired but reversible loss of erythrocyte complement receptor 1 (CR1, CD35) and its longitudinal alteration in patients with severe acute respiratory syndrome. Clin. Exp. Immunol. 139, 112–119.

Waxman, F. J., Hebert, L. A., Cornacoff, J. B., VanAman, M. E., Smead, W. L., Kraut, E. H. et al., 1984. Complement depletion accelerates the clearance of immune complexes from the circulation of primates. J. Clin. Investig. 74, 1329–1340.

Waxman, F. J., Hebert, L. A.,. Cosio, F. G., Smead, W. L., VanAman, M. E., Taguiam, J. M. et al., 1986. Differential binding of immunoglobulin A and immunoglobulin G1 immune complexes to pri-mate erythrocytes in vivo. Immunoglobulin A immune complexes bind less well to erythrocytes and are preferentially deposited in glomeruli. J. Clin. Investig. 77, 82–89.

Williamson, E., Walker, A.J., Bhaskaran, K., Bacon, S., Bates, C., Morton, C.E., et al., 2020. OpenSAFELY: factors associated with COVID-19-related hospital death in the linked electronic health records of 17 million adult NHS patients. https://doi.org/10.1101/2020.05.06.20092999

Wilson, J. G., Murphy, E. E., Wong, W. W., Klickstein, L. B., Weis, J. H., Fearon, D. T., 1986. Identification of a restriction fragment length polymorphism by a CR1 cDNA that correlates with the number of CR1 on erythrocytes. J. Exp. Med. 164, 50–59.

