## supplemental figures for "Acquired decrease of the C3b/C4b receptor (CR1, CD35) and increased C4d deposits on Erythrocytes from ICU COVID-19 Patients"

**Supplemental Figure 1. CR1/E density in COVID-19 patients according to days of disease.** (**A**), (**B**), (**C**), (**D**), CR1/E density patient followed-up were plotted with their longitudinal profile according to days of disease from the onset of symptoms in A, B, C, D chronological sub-figures for the sake of clarity. Thirty two COVID-19 patients are represented. Each is distinguished by one color and one symbol: circle, square, diamond, border, pentagon and hexagon.

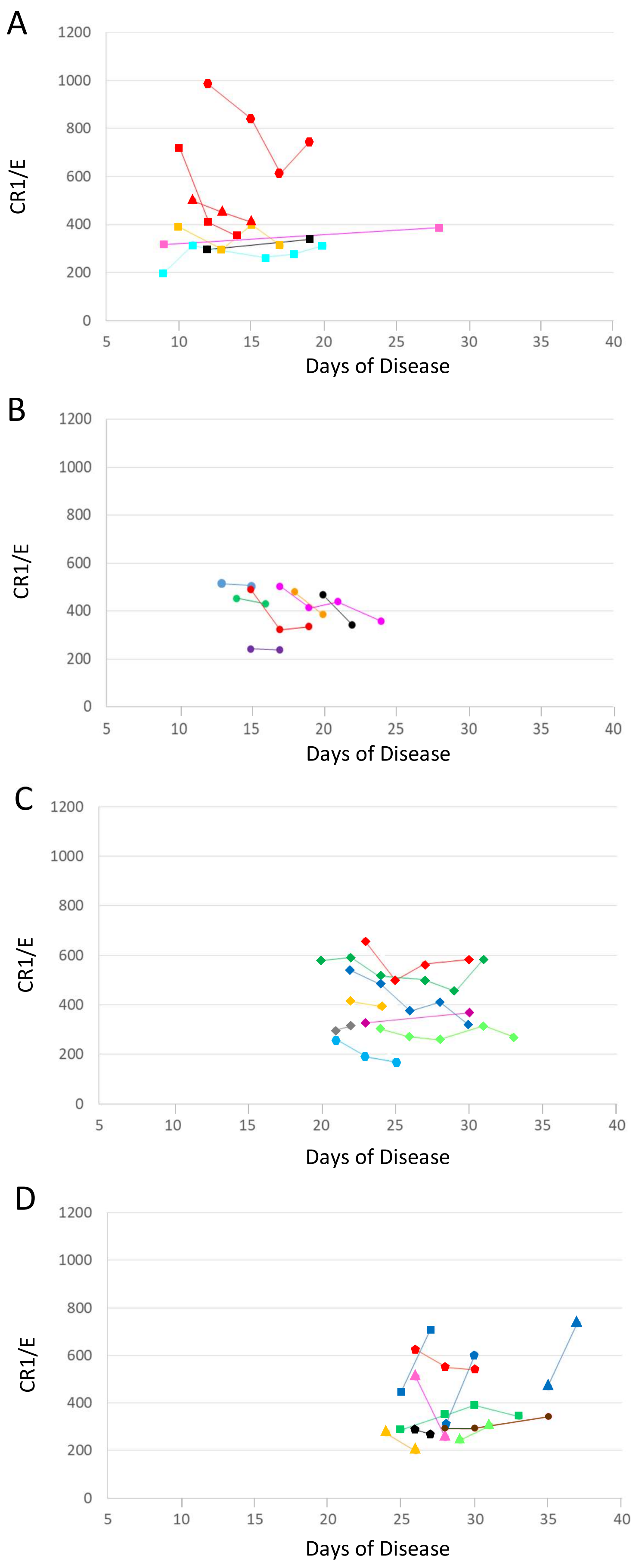

**Supplemental Figure 2. CR1/E density according to the arterial SaO2 decrease.** The CR1/E density is expressed as percentage of the CR1/E value predicted by the rs11118133 density genotype of the patient (CR1/E %). Mean decrease of CR1 for SaO2 under O2 (either optiflow or assisted ventilation) lower than 91 % was mean 49 % SD10 N14 versus mean 62 % SD10 N32 for SaO2 values above 91 % (p=0.007 Welch Two Sample t-Test). The lowest SaO2 value reached despite maximal O2 assistance was plotted against the CR1/E % at the first testing of each patient. Grey shadow SaO2 above 91%.
CR1/E values above 100 % originated either from the presence of reticulocytes which express 20000 more CR1 than E, or from a preferential destruction of old E in some haemolysis situations such as in septicemia.

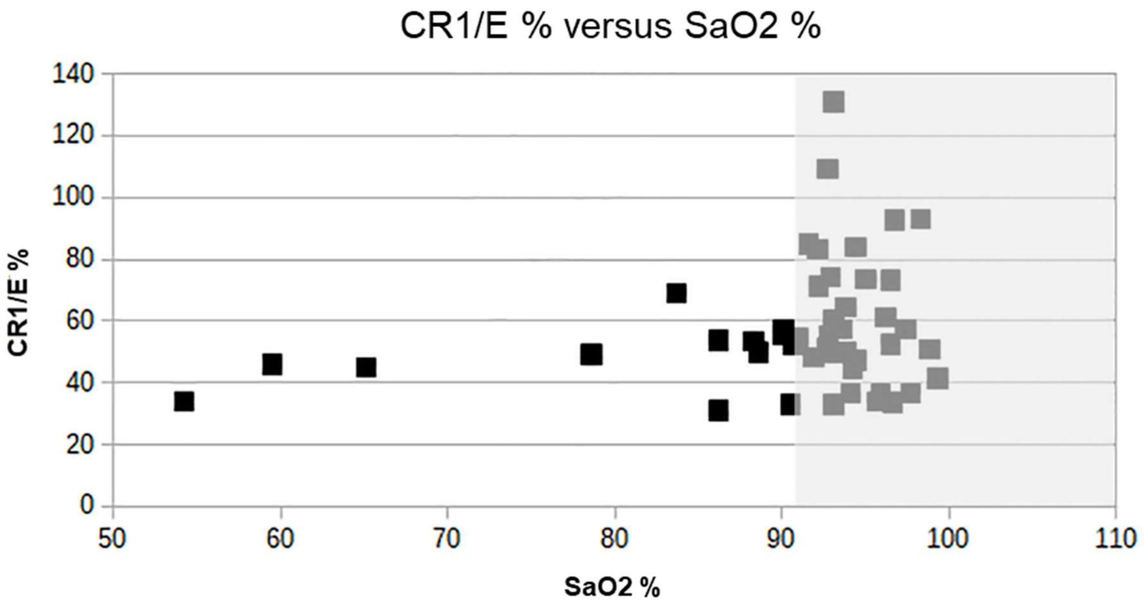


Supplemental material Figure 2

**Supplemental Figure 3. CR1/E density according to the level of D Dimers.** CR1/E density is expressed as percentage of the CR1/E value predicted by the rs11118133 density genotype of the patient (CR1/E %). The density of CR1/E tended to decrease as the level of D dimers increased.

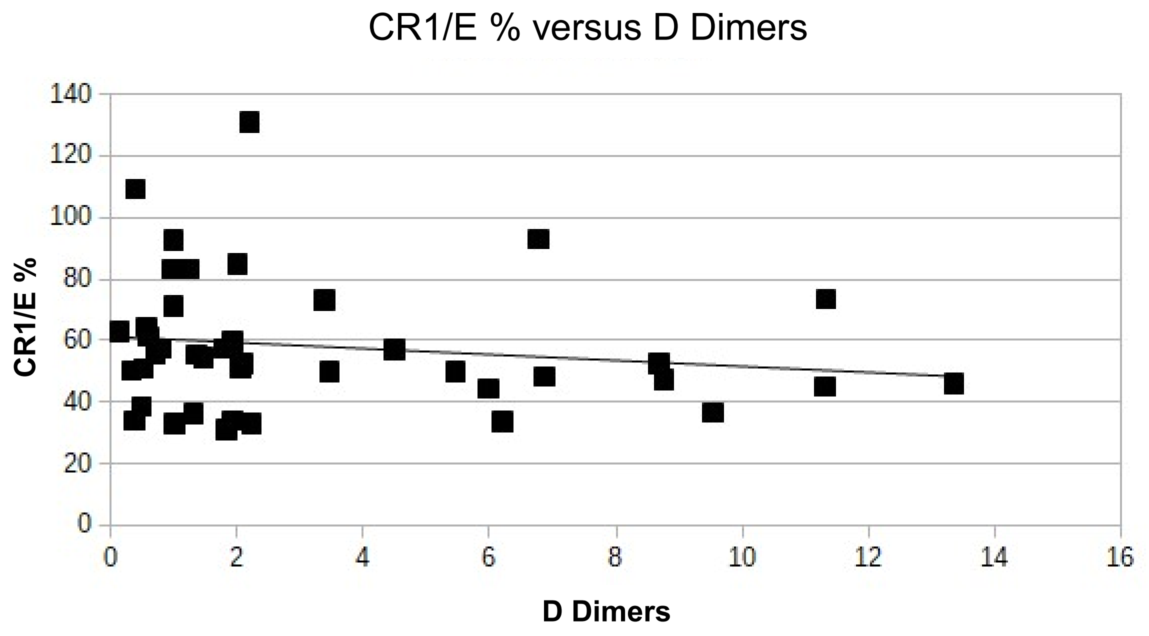


Supplemental material Figure 3

**Supplemental Figure 4. CR1/E density according to the dates of birth of the patients. (A**) The lowest measurement of CR1/E only was plotted for each patient according to the dates of birth. (**B**) The CR1/E density is expressed as percentage of the CR1/E value predicted by the rs11118133 density genotype of the patient (CR1/E %). Four patients didn't exhibit an acquired decrease of CR1/E below the density predicted by their genotypes.

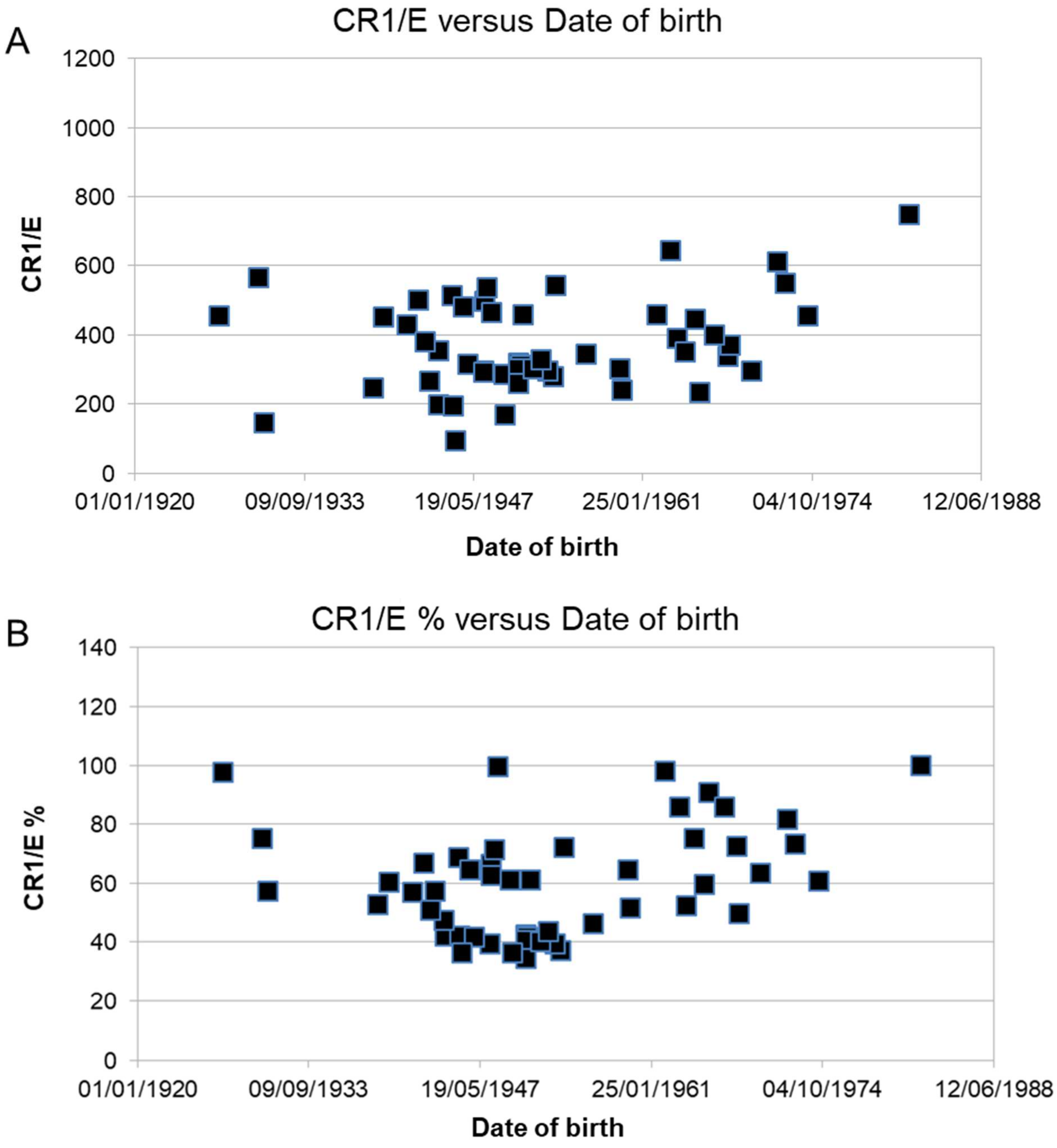


Supplemental material Figure 4

**Supplemental Figure 5. CR1/E density among deceased patients according to the duration of the disease, from the first day of symptoms to the day of blood collection.** CR1/E is expressed as percentage of the mean CR1/E density expected from the rs11118133 density genotype of a given individual (CR1/E %).
Triangle: homozygous LL low density genotype. Circle: heterozygous HL medium density genotype. Square: homozygous HH high density genotype.
Among the 10 deceased patients with globally low CR1/E values, the earlier in the disease the CR1/E was assessed, the lower were the CR1/E values. The equation of the line obtained is f(x) = 0.856 x + 42.80 $R^2$ = 0.696.

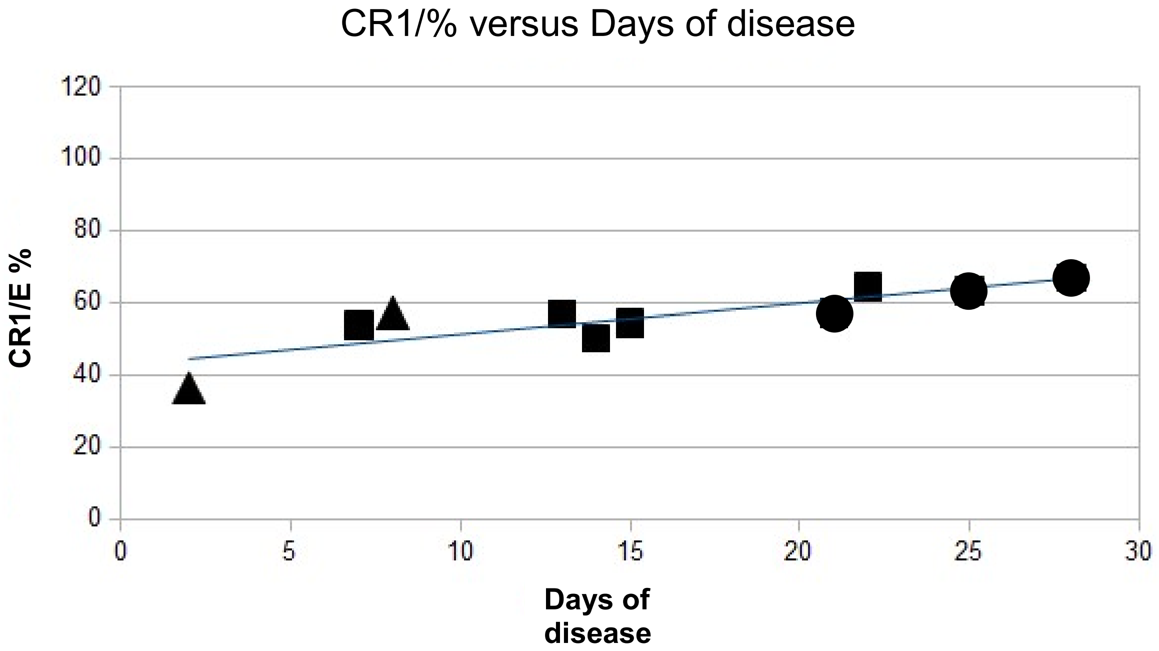


Supplemental material Figure 5

**Supplemental Figure 6. CR1/E density according to the C4d deposits.** The COVID-19 patients are depicted in black. The healthy controls are depicted in red. X axis: The C4d deposits on E are expressed as positive events above the negative control staining.

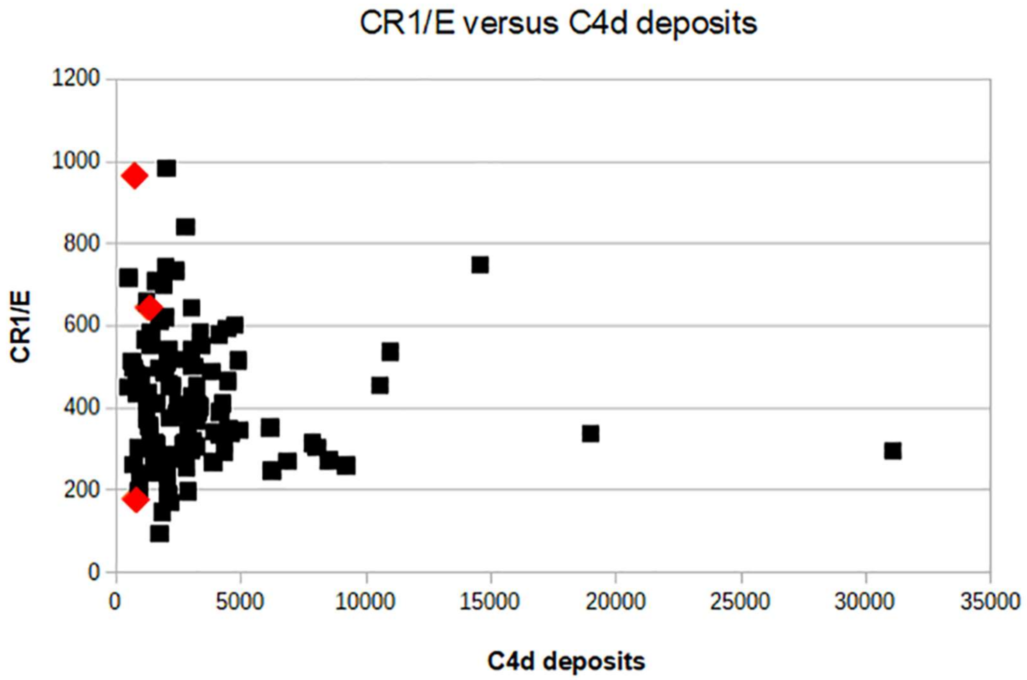


Supplemental material Figure 6
